# Integrated monogenic and polygenic risk predicts disease progression in Fuchs endothelial corneal dystrophy

**DOI:** 10.64898/2026.02.17.26346339

**Authors:** Siyin Liu, Anita Szabo, Christina Zarouchlioti, Nihar Bhattacharyya, Quang Nguyen, Marcos Abreu Costa, Robert N Luben, Lubica Dudakova, Pavlina Skalicka, Martin Horak, Anthony P. Khawaja, Nikolas Pontikos, Kirithika Muthusamy, Stephen J. Tuft, Petra Liskova, Alice E. Davidson

**Affiliations:** UCL Institute of Ophthalmology, London, United Kingdom; Moorfields Eye Hospital, London, United Kingdom; Department of Paediatrics and Inherited Metabolic Disorders, First Faculty of Medicine, Charles University and General University Hospital in Prague, Prague, Czech Republic; Department of Ophthalmology, First Faculty of Medicine, Charles University and General University Hospital in Prague, Prague, Czech Republic; Department of Ophthalmology, University Hospital Královské Vinohrady and 3rd Faculty of Medicine, Charles University, 100 34, Prague 10, Czech Republic; NIHR Biomedical Research Centre, Moorfields Eye Hospital NHS Foundation Trust & UCL Institute of Ophthalmology, London, UK

**Keywords:** Fuchs endothelial corneal dystrophy, polygenic risk score, CTG18.1

## Abstract

**Purpose:** Fuchs endothelial corneal dystrophy (FECD) is a common corneal disease and a leading indication for endothelial keratoplasty (EK). Although CTG18.1 repeat expansion is a major genetic risk factor, the contribution of polygenic background to disease progression remains unclear. We evaluated whether combining CTG18.1 expansion status with a FECD-specific polygenic risk score (PRS) enables genomic prediction of progression to EK.

**Methods:** We retrospectively analysed 589 individuals with FECD from two European centers, with replication in an independent cohort of 185 individuals. Association of CTG18.1 expansion (≥50 repeats) and PRS with time to EK were evaluated using Cox models adjusted for sex and ancestry.

**Results:** Expansion-positive status was associated with earlier EK (HR 2.30; 95% CI 1.62– 3.26; P<.001). Addition of PRS improved prediction (C-index 0.614 vs 0.602; P=.014). Each 1-SD increase in PRS was associated with earlier EK (HR 1.16; 95% CI 1.03–1.30; P=.015), with replication in the validation cohort (HR 1.42; 95% CI 1.15–1.75; P=.001).

**Conclusion:** Integration of monogenic and polygenic risk enables genomic prediction of FECD progression, supporting clinical genomic risk stratification to inform individualized monitoring and timing of intervention.

## Introduction

Fuchs endothelial corneal dystrophy (FECD) is a common inherited disorder characterized by progressive corneal endothelial failure and visual impairment.^1,2^ Although endothelial keratoplasty (EK) remains the definitive intervention, progression to keratoplasty is unpredictable, as many individuals never require surgery, whereas others progress rapidly to end-stage disease. Consequently, predicting risk and timing of progression to surgical intervention remains a major clinical challenge. At present, clinicians lack a validated prognostic framework. With several non-surgical and disease-modifying therapies entering clinical evaluation,^1,3,4^ the need for reliable markers to identify patients most likely to progress is pertinent.

The CTG18.1 trinucleotide repeat expansion is the dominant risk for FECD in Europeans and South Asians.^5^ Large genome-wide association studies (GWAS) have also uncovered a multiple risk loci,^6,7^ suggesting that polygenic background shapes disease risk.

Polygenic risk scores (PRS) have improved risk prediction and demonstrated prognostic utility, including in several complex ophthalmic disorders.^8–11^ There is also evidence that PRS can distinguish individuals with FECD from unaffected controls,^6,7^ but its utility for predicting disease progression is untested. Here, we evaluate whether integration of monogenic and polygenic risk supports genomic prediction of clinical progression in FECD.

## Methods

We retrospectively identified 589 unrelated individuals of genetically confirmed European ancestry in a multicenter genotype-phenotype study of clinically confirmed FECD,^5^ with a validation cohort of 185 patients **(Supplementary Method; Supplementary Figure 1**). We excluded individuals with prior intraocular surgery. The primary outcome was age at first EK in either eye, and individuals without surgery were censored at their last follow-up visit. Institutional review boards approved the study (22/PR/0482; 22/EE/0090; 13/LO/1084; 42/24), and all participants provided written informed consent.

Genome-wide genotyping of the primary and validation cohorts was performed using the UK Biobank Axiom Array, with standard variant and sample quality-control filters (**Supplementary Method**). Imputation used the TOPMed GRCh38 reference panel.

CTG18.1 repeat status was determined using short tandem repeat-PCR and triplet-primed PCR assays^12^: Individuals with one or both alleles ≥50 repeats were categorised as expansion-positive (Exp+) and those with biallelic alleles of <50 repeats as expansion-negative (Exp-).

PRS were constructed using PRSice-2^13^ from an independent European FECD GWAS summary statistics^14^ **(Supplementary Methods)**. PRS were standardized as Z-scores (mean 0, SD 1).

Analyses were performed in R (version 4.3.1). Associations between PRS and time to keratoplasty were evaluated using Cox proportional hazards regression adjusted for sex, CTG18.1 expansion status, and ancestry principal components. Hazard ratios were expressed per 1-SD increase in PRS. Model discrimination was assessed using Harrell’s C-index, and transplant-free survival across PRS quartiles was visualised with Kaplan-Meier curves. A two-sided P < .05 was considered statistically significant (**Supplementary Method)**.

## Results

The primary cohort included 589 individuals with FECD; 365 (62%) female, 456 (77%) CTG18.1 Exp+, and 347 (59%) had undergone EK (mean [SD] age at surgery, 68 [9] years). In the validation cohort, 117 of 185 (63%) were female, 144 (78%) were Exp+, and 120 (65%) had an EK. Baseline characteristics by PRS quartile are shown in **Supplementary**

**Tables 1 and 2**. In both cohorts the mean (SD) PRS was higher among participants who underwent EK (primary cohort: 0.12 ± 1.05 versus −0.20 ± 0.94, P < .001; validation cohort: 0.20 ± 0.95 versus −0.29 ± 1.01, P = .001; **Supplementary Figure 2)**.

**Table 1.** Comparison of Cox Proportional Hazards Models for Predicting Time to Keratoplasty in the Primary Cohort (N =589)

| **Model / Subgroup** | **Predictor** | **Hazard Ratio**<br>**(95% CI)** | **P Value** | **Concordance**<br>**(SE)** |
| --- | --- | --- | --- | --- |
| **Base model**<br>**(Overall)** | CTG18.1<br>expansion (Exp+) | 2.30 (1.62–3.26) | < .001 | 0.602 (0.017) |
|  | Sex (male) | 1.08 (0.87–1.35) | .48 |  |
| **Full model**<br>**(Overall)** | CTG18.1<br>expansion (Exp+) | 2.15 (1.51–3.05) | < .001 | 0.614 (0.017) |
|  | PRS (per 1-SD<br>increase) | 1.16 (1.03–1.30) | .015 |  |
|  | Sex (male) | 1.07 (0.85–1.34) | .57 |  |
| *Base model includes CTG18.1 expansion status, sex, and 10 ancestry principal components. Sex (male) is modelled relative to female (reference category). Full model additionally includes standardized polygenic risk score (PRS). |  |  |  |  |

There was a progressive increase in the likelihood of EK with a higher PRS. The proportion with EK increased across PRS deciles (Cochran-Armitage Z = 3.69; P < .001; **Figure 1A**), and individuals in the top decile underwent surgery at younger ages than those in the lowest (mean, 64.6 vs 70.1 years; P = .005; **Figure 1B**). The odds of EK increased progressively across the PRS thresholds, reaching 3.20-fold higher odds in the top 10% compared with the remainder (95% CI, 1.65-6.58; P < .001; **Supplementary Table 3**).

**Figure. 1.**
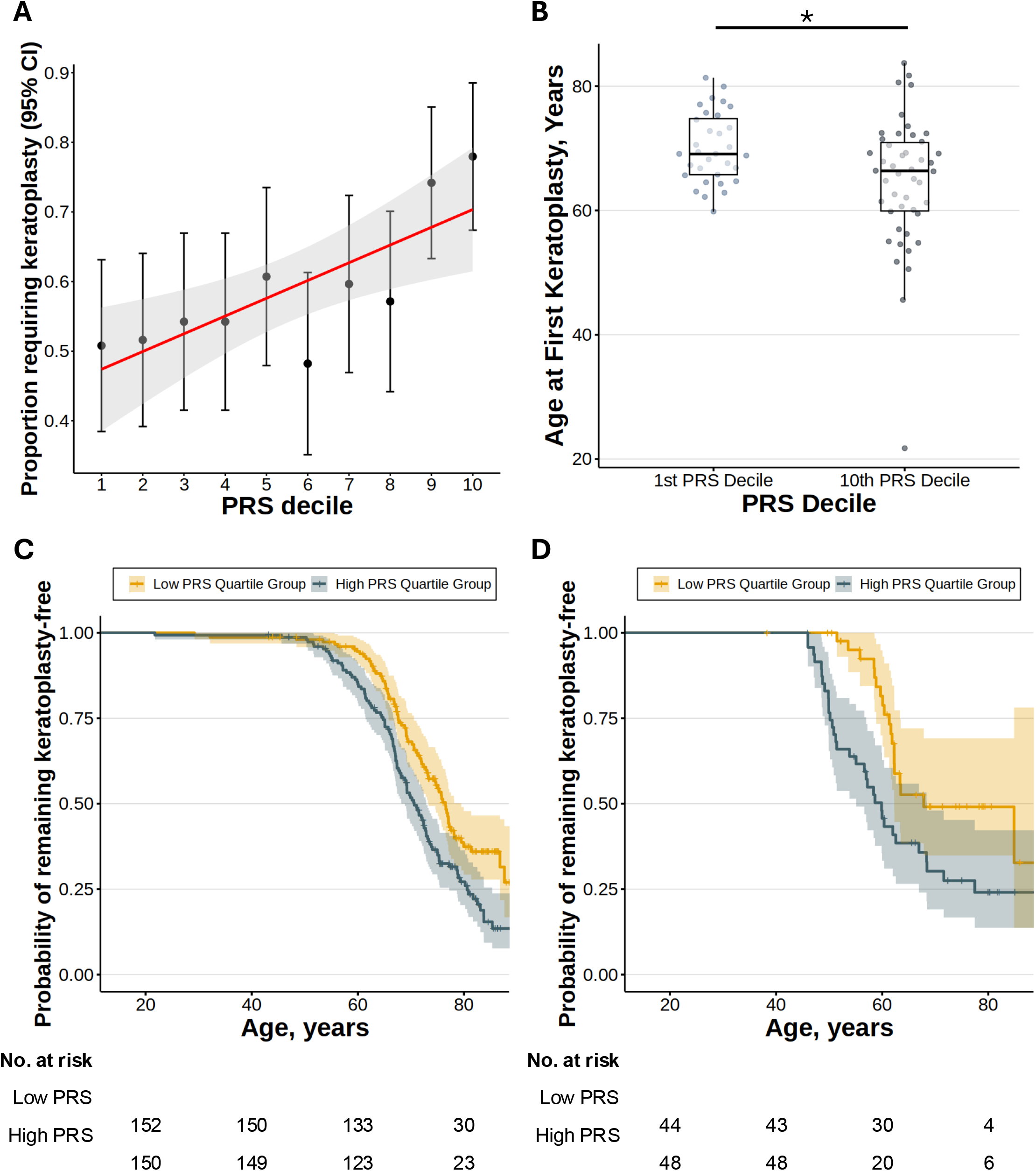
Polygenic burden is associated with increased and earlier keratoplasty in FECD. (A) The proportion with EK increased across polygenic risk score (PRS) deciles, demonstrating a significant dose-response trend (Z = 3.69; P < .001). (B) Patients in the highest PRS decile had a first EK at a younger age (mean 64.6 ± 10.6 years) compared with those in the lowest decile (70.1 ± 5.6 years; mean difference = 5.4 years; P = .005). (C) Kaplan–Meier curves of EK-free survival in the primary cohort stratified by PRS quartiles show significantly earlier progression in the highest versus lowest quartile (log-rank P = .0008). (D) The association between higher PRS and earlier EK was replicated in an independent validation cohort (log-rank P = .008). Shaded areas represent 95% confidence intervals.

After adjustment for sex and ancestry principal components, Exp+ status was strongly associated with earlier progression to EK (HR, 2.30; 95% CI, 1.62–3.26; P < .001; **Table 1**). To determine whether polygenic burden provided additional prognostic information, the PRS was added to the model. Each 1-SD increase in PRS conferred 16% higher hazard of EK (HR, 1.16; 95% CI, 1.03–1.30; P = .015). When comparing these two nested models, the PRS Z-score improved discrimination performance (C-index, full model 0.614 vs base model 0.602; likelihood-ratio χ^2^ = 6.04; P = .014; **Table 1**), and yielded consistently higher time-dependent AUC(t) values (Wilcoxon signed-rank V = 1; P = .03; **Supplementary Figure 3**). Sensitivity analyses stratified by CTG18.1 expansion status demonstrated that higher PRS remained associated with time to EK in both Exp+ and Exp− subgroups (**Supplementary Results**).

Absolute risk estimates demonstrated the clinical impact of polygenic burden (**Figure 1C)**. By age 60 years, the cumulative incidence of EK was 5.4% in the lowest PRS quartile compared with 15.1% in the highest quartile (2.76-fold increase, 9.6% absolute difference [95% CI 2.7–16.5%]; P = .011); by age 70 years, 31.9% versus 48.3% (1.51-fold increase, 16.4% absolute difference [95% CI 4.9–27.8%]; P = .002); and by age 80, 62.6% versus 72.8% (1.16-fold increase, 10.2% absolute difference [95% CI −2.0–22.5%]; P = .005).

These findings were replicated in the validation cohort (**Figure 1D)**, where each 1 SD increase in PRS was associated with a 42% higher hazard of keratoplasty (HR, 1.42; 95% CI, 1.15–1.75; P = .001; **Supplementary Table 6**). Full dose-response analyses and subgroup results are provided in **Supplementary Results**.

## Discussion

To the best of our knowledge, this is the first study to demonstrate that both CTG18.1 expansion status and polygenic burden can be used to quantify the risk for EK in FECD. CTG18.1 expansion, the major known genetic risk factor for FECD,^5^ was associated with earlier progression. Importantly, inclusion of a FECD-specific PRS provided enhanced prognostic stratification beyond repeat expansion status alone. These findings were validated in an independent cohort.

The progressive increase in surgical risk across the PRS distribution suggests that the cumulative burden of common variants contributes to heterogeneity in progression. This parallels other complex traits where monogenic and polygenic factors jointly influence clinical expression.^15,16^ Our findings therefore support a liability-threshold model where a high-penetrance repeat expansion and polygenic background collectively modify disease progression. The stronger predictive value of PRS among Exp-individuals also suggests that common variant burden likely plays a more pronounced role in the absence of the expansions. Together, these results support a gradation of FECD genetic risk.

FECD associated with CTG18.1 expansion occurs predominantly in individuals of European ancestry, and the largest available discovery GWAS datasets are European. Accordingly, analyses were restricted to genetically confirmed European ancestry to minimize population stratification and optimize PRS calibration. Evaluation of PRS performance across diverse ancestral groups is essential for equitable clinical translation and is a priority for future studies.

At present, no established clinical or genetic prediction model exists for FECD progression. This genomic-based model showed consistent stratification of absolute risk at clinically relevant age range. Genetic profiling could therefore facilitate individualised counselling at diagnosis regarding the likelihood and timing of future keratoplasty and assist prioritization for closer follow-up in resource-limited settings. For example, individuals in higher PRS strata could reasonably be considered for closer surveillance to detect early decompensation, and patients with early FECD undergoing cataract surgery may benefit from tailored surgical approaches to minimize endothelial stress. In addition, genomic-informed stratification may be valuable for clinical trials of emerging disease-modifying therapies, where enriching enrollment for individuals at higher near-term risk of progression could improve efficiency and reduce required sample sizes.

Given the contributions of environmental influences, including oxidative stress and sex, robust prediction for disease progression based on PRS alone is unlikely. However, with further refinement in larger GWASs to identify more risk loci, and incorporating non-genetic predictors, PRS-based risk profiling could inform care pathways and refine patient selection for clinical trials. Consistent with recent ACMG recommendations, clinical application of PRS into practice will require prospective validation, evaluation across diverse populations, and integration with established clinical predictors.^17^ PRS should be considered a complementary component, not a standalone determinant of care.

### Limitations

Although we used the largest available genetically characterized FECD datasets (targeted CTG18.1 and genome-wide SNP array), the relatively modest sample size constrains precision in subgroup estimates. The European ancestry of the cohorts restricts the generalizability of our findings. Using the time to EK as the endpoint, although clinically relevant, is influenced by healthcare access and surgical practice.

## Conclusions

This study underpins the value of genetic profiling for FECD prognosis, defined as the risk of EK, which provides a foundation for genetically informed risk stratification of FECD progression. As cost-effective genotyping becomes routine and disease-modifying therapies advance, validated PRS-informed risk assessment may enable the identification of individuals at greatest risk of progression, facilitating earlier and targeted interventions to prevent vision loss.

## Data Availability

In this study we used patient identifiable information. For this reason raw data cannot be made publicly available due to confidentiality considerations.

## Acknowledgments

The authors thank the patients and their families for their participation and support. The authors are grateful to the clinical staff at Moorfields Eye Hospital and General University Hospital and Beverly Scott for technical support. We thank Jana Jedlickova and Beverly Scott for their technical support.

## Funding/Support

This work was funded by a UKRI Future Leader Fellowship MR/S031820/1 and MR/Y019911/1 (A.E.D.), Medical Research Council (S.L and A.E.D, MR/X006271/1), Moorfields Eye Charity GR001395 (A.E.D.), Fight for Sight 5171 / 5172 (A.E.D.), Sight Research UK SEE027 (S.L). The National Institute for Health Research Biomedical Research Centre at Moorfields Eye Hospital National Health Service Foundation Trust and University College London Institute of Ophthalmology NIHR203322. APK is supported by a UK Research and Innovation Future Leaders Fellowship (MR/Y033930/1), an Alcon Research Institute Young Investigator Award and a Lister Institute for Preventive Medicine Award. P.L., L.D., and P.S. were supported by the Ministry of Health of the Czech Republic (NW25-07-00303) and Univerzita Karlova v Praze (UNCE/24/MED/022 and SVV 2600631).

## Author contributions

Concept and design: Liu, Davidson

Data acquisition and analysis: Liu, Szabo, Zarouchlioti, Bhattacharyya, Nguyen, Costa, Skalicka, Liskova, Dudakova, Horak

Drafting of the manuscript: Liu, Davidson

Critical review of the manuscript for important intellectual content: All authors

Statistical analysis: Liu, Luben, Pontikos

Obtained funding: Liu, Liskova, Davidson, Dudakova

Supervision: Davidson

## Conflict of Interest Disclosures

Dr Davidson reported grants from the UK Research and Innovation Future Leader Fellowship, the Medical Research Council, Moorfields Eye Charity, Sight Research UK, Fight for Sight, and the Rosetrees Trust during the conduct of the study; research collaboration with Prime Medicine. No other disclosures were reported. APK has acted as a paid consultant or lecturer to Abbvie, Aerie, Google Health, Heidelberg Engineering, Glaucore, Novartis, Qlaris Bio, Regeneron, Reichert, Santen, Seonix Bio, Thea and Topcon.

## Ethics Declaration

The study was approved by the relevant institutional research ethics committees at participating centers (22/PR/0482; 22/EE/0090; 13/LO/1084; 42/24). All participants provided written informed consent. The study was conducted in accordance with the Declaration of Helsinki.

## Notes

### Author Declarations

The study was approved by the Research Ethics Committees of University College London (UCL) (22/EE/0090), Moorfields Eye Hospital (13/LO/1084), and the General University Hospital (151/11 S-IV). All participants provided written informed consent. The study was conducted in accordance with the Declaration of Helsinki.

