## Supplemental Materials for "Integrated monogenic and polygenic risk predicts disease progression in Fuchs endothelial corneal dystrophy"

### Supplementary Method

##### Study Population and Genome-Wide Array Genotyping

Two independent cohorts of individuals with clinically confirmed Fuchs endothelial corneal dystrophy (FECD) and genetically confirmed European ancestry recruited from Moorfields Eye Hospital, London, and the General University Hospital, Prague were analyzed. The primary cohort were recruited as part of ongoing genetic studies of FECD[1]. An independent, non-overlapping validation cohort from the same sites was recruited using identical protocols. All participants were diagnosed with FECD based on the documented finding of confluent corneal guttae seen by slit-lamp examination. They provided written informed consent and whole blood or saliva for DNA extraction. Genomic DNA was extracted from whole blood or saliva using a Gentra Puregene Blood kit (Qiagen) or Oragene saliva kit (Oragene OG-300, DNA Genotek). To prevent the confounding effect of traumatic endothelial cell loss, patients with a history of intraocular surgery, including cataract extraction, were excluded. Similarly, we only included patients who had a primary endothelial keratoplasty with or without planned phacoemulsification. The primary outcome was time from birth to the first endothelial keratoplasty; participants without keratoplasty were censored at last clinical follow-up. This study followed Strengthening the Reporting of Observational Studies in Epidemiology cohort study guidelines.

Genomic DNA was extracted from peripheral blood or saliva using standard protocols. Genotyping was performed with the UK Biobank Axiom array (Axiom UKB WCSG.r5; Thermo Fisher Scientific) at the Ramaciotti Centre for Genomics, Sydney, Australia. The genotypes were called using Applied Biosystem Analysis Power Tools.[2] The recommended filters were used in the quality control steps. Generic priors were used as advised in the user manual for larger batches (>96 samples). The genotyped dataset was filtered by PLINK2[3] based on missingness per individual (> 0.05), missingness per marker (> 0.05) and minor allele frequency (< 0.01). TOPMed reference panel (topmed-r3@2.0.0-beta3, hg38) was used, and the imputation was performed on TOPMed Imputation Server.[4] The original hg19 human genome build was lifted to hg38, phased by Eagle v2.4[5] and imputed by Minimac4.[6] SNPs with a $r^2$ (imputation quality) <0.2 were excluded.

Post-imputation QC removed multiallelic variants, duplicate variant IDs, variants with MAF < 0.05, imputation quality $r^2 < 0.8$, or genotype missingness > 0.02. Only high-quality autosomal variants were retained for polygenic analyses. Only individuals of genetically confirmed European ancestry, confirmed by principal component analysis (FRAPOSA)[7], were included to minimize population stratification.

##### CTG18.1 Expansion Genotyping

The CTG18.1 trinucleotide repeat expansion in *TCF4* was genotyped using a previously-described short tandem repeat (STR)-polymerase chain reaction (PCR) assay[8]. Triplet repeat-primed (TP)-PCR was subsequently performed if only one CTG18.1 allele was detected, to determine if an allele longer than the STR-PCR detection maximum (~125 repeats) was present. Amplicons were analysed using capillary electrophoresis, and repeat length was determined using GeneMapper software (Applied Biosystems). Individuals with one or more expanded alleles (≥50 CTG repeats) were classified as expansion-positive (Exp+); all others were Exp-.

**Polygenic Risk Score Derivation**

Polygenic risk scores (PRSs) were calculated via a clumping-and-thresholding approach using PRSice-2 software.[9] Base SNP weights were obtained from the largest available FECD GWAS (3,473 cases and 445,239 controls of European ancestry).[10] The current study cohorts were not included in this GWAS. The SNPs that overlapped between the summary statistics GWASs (base dataset) and local SNP dataset (target dataset) were extracted.

Linkage disequilibrium (LD) clumping was carried out in PRSice-2 using a 500-kb window and $r^2 < 0.1$. SNPs with minor allele frequency <1% or genotype missingness >1% were excluded. PRSs were calculated across a series of P-value thresholds ($5\times10^{-8}$ to0.5). The top 10 ancestry principal components (PC) derived from the target genotype dataset were included in all downstream analyses.

In the primary cohort, the optimal P-value threshold was identified by maximising Harrell's C-index in a Cox proportional hazards model for time to first keratoplasty, adjusting for sex, CTG18.1 expansion status, and ancestry PCs. In the validation cohort, PRSs were computed using the same SNP subset and effect-size weights derived from the primary cohort without further tuning. All PRSs were standardised to Z-scores (mean = 0, SD = 1).

**Statistical Analysis**

The primary analysis evaluated the association between PRS Z-scores and time to keratoplasty using Cox proportional hazards regression (coxph function, survival package in R v4.3.1). Age was the underlying time scale. The base model included sex and the top 10 PCs. The full model additionally included CTG18.1 expansion status. Hazard ratios (HRs) were expressed per 1-SD increase in PRS.

Model discrimination was quantified using Harrell's concordance index (C-index). Improvement in model fit after adding PRS Z-scores was tested using the likelihood-ratio test. Time-dependent AUC(t) was computed at 5-year intervals, and mean differences between the base and full models were compared using the Wilcoxon signed-rank test.

Transplant-free survival across PRS quartiles was estimated using Kaplan-Meier curves, with differences assessed by log-rank testing. Dose-response associations across PRS deciles were evaluated using the Cochran-Armitage trend test and linear regression of the proportion censored against PRS decile.

Continuous variables were compared across cohorts using t-tests or one-way ANOVA, as appropriate. Categorical variables were compared using $\chi^2$ tests or Fisher's exact tests when cell counts were small. Distributions were summarised as mean (SD) or median (IQR) for continuous variables and as counts (%) for categorical variables.

Sensitivity analyses were performed in the following sub-groups to assess robustness:

1. CTG18.1 expansion-positive subgroup (Exp+): To test whether PRS contributes additively beyond the monogenic repeat expansion.
2. CTG18.1 expansion-negative subgroup (Exp-): To quantify polygenic contribution in the absence of the expansion.

All tests were two-sided, with $P < .05$ considered statistically significant.

### Supplementary Results

**Primary Cohort Sensitivity Analysis Results**
In a sensitivity analysis restricted to Exp+ participants only, higher PRS remained significantly associated with time to keratoplasty, with each 1-SD increase in PRS conferred a 13.7% higher hazard of keratoplasty (HR 1.14; 95% CI 1.01–1.29; P = .039), indicating an additive effect of common polygenic variation beyond the repeat expansion (**Supplementary Table 4)**. Among Exp- participants, where the expansion was absent, each 1-SD increase in PRS was also associated with a 51% higher hazard of keratoplasty (HR 1.51; 95%CI 1.00–2.28; P = .048), suggesting that polygenic background contributes substantially to disease progression in individuals without the major effect from the CTG18.1 expansion (**Supplementary Table 4)**.

**Validation Cohort Results**
Findings in the validation cohort were directionally consistent with the primary cohort. Individuals in the top 10% of the PRS distribution had 6.39-fold higher odds of requiring keratoplasty (95% CI, 1.65–42.63; P = .02; **Supplementary Table 5**) compared with the remainder of the cohort**.** The addition of the PRS improved overall model performance in the validation cohort, with the C-index increasing from 0.679 (SE = 0.026) in the baseline model to 0.689 (SE = 0.026) in the full model (likelihood-ratio test $\chi^2 = 11.17$; df = 1; P < .001; **Supplementary Table 6**). Sensitivity analyses in the validation cohort reproduced the pattern observed in the primary cohort, with PRS remaining significantly associated with keratoplasty risk among both Exp+ (HR 1.40; 95% CI, 1.11–1.75; P = .005) and Exp- individuals (HR 2.94; 95% CI, 1.13–7.68; P = .028), supporting the robustness of the PRS effect across genetic subgroups (**Supplementary Table 6)**.

### Supplementary Tables

**Supplementary Table 1. Summary of the Primary Cohort and Comparison of Baseline Characteristics in Polygenic Risk Score (PRS) Risk Quartiles**

| Characteristics | Overall (N = 589) | PRS Risk Quartile | | | | P value |
|---|---|---|---|---|---|---|
| | | Quartile 1 (N = 152) | Quartile 2 (N = 145) | Quartile 3 (N = 142) | Quartile 4 (N = 150) | |
| **Sex** | | | | | | .11 |
| Female | 365 (62%) | 97 (64%) | 100 (69%) | 79 (56%) | 89 (59%) | |
| Male | 224 (38%) | 55 (36%) | 45 (31%) | 63 (44%) | 61 (41%) | |
| **CTG18.1 Expansion Status** | 456 (77%) | 99 (65%) | 108 (74%) | 111 (78%) | 138 (92%) | < .001 |
| Biallelic expanded cases | 33 (5.6%) | 2 (1.3%) | 6 (4.1%) | 8 (5.6%) | 17 (11%) | |
| Monoallelic expanded cases | 423 (72%) | 97 (64%) | 102 (70%) | 103 (73%) | 121 (81%) | |
| Non-expanded | 133 (23%) | 53 (35%) | 37 (26%) | 31 (22%) | 12 (8.0%) | |
| **Endothelial keratoplasty status** | | | | | | .002 |
| Had keratoplasty | 347 (59%) | 78 (51%) | 83 (57%) | 78 (55%) | 108 (72%) | |
| No keratoplasty | 242 (41%) | 74 (49%) | 62 (43%) | 64 (45%) | 42 (28%) | |
| **Survival time (mean [SD])** | 71 (11) | 71 (11) | 72 (11) | 73 (11) | 69 (11) | .04 |
| **Age at first keratoplasty (among cases with keratoplasty)** | 68 (9) | 68 (9) | 69 (8) | 70 (9) | 67 (9) | .14 |

**Supplementary Table 2. Summary of the Validation Cohort and Comparison of Baseline Characteristics in Polygenic Risk Score (PRS) Risk Quartiles**

| **Characteristics** | **Overall (N = 186)** | **PRS Risk Quartile** | | | | **P value** |
|---|---|---|---|---|---|---|
| **Sex** | | **Quartile 1 (N = 44)** | **Quartile 2 (N = 45)** | **Quartile 3 (N = 49)** | **Quartile 4 (N = 48)** | |
| Female | 117 (63%) | 29 (66%) | 27 (60%) | 35 (71%) | 26 (54%) | .3 |
| Male | 69 (37%) | 15 (34%) | 18 (40%) | 14 (29%) | 22 (46%) | |
| **CTG18.1 Expansion Status** | 144 (78%) | 29 (66%) | 31 (69%) | 38 (78%) | 46 (96%) | .002 |
| Biallelic expanded cases | 6 (3.2%) | 0 (0%) | 0 (0%) | 4 (8.2%) | 2 (4.2%) | |
| Monoallelic expanded cases | 138 (74%) | 29 (66%) | 31 (69%) | 34 (69%) | 44 (92%) | |
| Non-expanded | 42 (23%) | 15 (34%) | 14 (31%) | 11 (22%) | 2 (4.2%) | |
| **Endothelial keratoplasty status** | | | | | | .009 |
| Had keratoplasty | 120 (65%) | 19 (43%) | 32 (71%) | 36 (73%) | 33 (69%) | |
| No keratoplasty | 66 (35%) | 25 (57%) | 13 (29%) | 13 (27%) | 15 (31%) | |
| **Survival time (mean [SD])** | 62 (13) | 65 (11) | 62 (11) | 59 (14) | 61 (14) | .2 |
| **Age at first keratoplasty (among cases with keratoplasty)** | 58 (8) | 61 (7) | 60 (9) | 56 (9) | 56 (8) | .018 |

**Supplementary Table 3. Odds of Keratoplasty According to Increasing Polygenic Risk Distribution in the Primary Cohort**

| PRS distribution | Reference | OR (95% CI) | p-value |
|---|---|---|---|
| Top 50% of distribution | Remaining 50% | 1.44 (1.00–2.08) | $5.09 \times 10^{-2}$ |
| Top 40% of distribution | Remaining 60% | 1.74 (1.20–2.55) | $3.76 \times 10^{-3}$ |
| Top 30% of distribution | Remaining 70% | 2.00 (1.33–3.04) | $9.09 \times 10^{-4}$ |
| Top 20% of distribution | Remaining 80% | 2.80 (1.72–4.66) | $5.12 \times 10^{-5}$ |
| Top 10% of distribution | Remaining 90% | 3.20 (1.65–6.58) | $8.93 \times 10^{-4}$ |

**Supplementary Table 4. Comparison of Base and Full Cox Proportional Hazards Models for Predicting Time to Keratoplasty in the Primary Cohort**

| Model / Subgroup | Predictor | Hazard Ratio (95% CI) | P Value | Concordance (SE) |
|---|---|---|---|---|
| **Base model (Overall)** | CTG18.1 expansion (Exp+) | 2.30 (1.62–3.26) | < .001 | 0.602 (0.017) |
| | Sex (male) | 1.08 (0.87–1.35) | .48 | |
| **Full model (Overall)** | CTG18.1 expansion (Exp+) | 2.15 (1.51–3.05) | < .001 | 0.614 (0.017) |
| | PRS (per 1-SD increase) | 1.16 (1.03–1.30) | .015 | |
| | Sex (male) | 1.07 (0.85–1.34) | .57 | |
| Sensitivity Analysis | | | | |
| **Full model (CTG18.1 Exp+ only)** | PRS (per 1-SD increase) | 1.14 (1.01–1.29) | .04 | 0.569 (0.018) |
| | Sex (male) | 1.10 (0.87–1.39) | .44 | |
| **Full model (CTG18.1 Exp- only)** | PRS (per 1-SD increase) | 1.51 (1.00–2.28) | .0475 | 0.799 (0.037) |
| | Sex (male) | 1.44 (0.55–3.79) | 0.4558 | |
| *Base model includes CTG18.1 expansion, sex, and 10 genetic principal components (PCs). Full model additionally includes standardized polygenic risk score (PRS). | | | | |

**Supplementary Table 5. Odds of Keratoplasty According to Increasing Polygenic Risk Distribution in the Validation Cohort**

| PRS distribution | Reference | OR (95% CI) | P value |
|---|---|---|---|
| Top 50% of distribution | Remaining 50% | 1.42 (0.74–2.76) | $2.95x10^{-1}$ |
| Top 40% of distribution | Remaining 60% | 1.59 (0.82–3.14) | $1.76x10^{-01}$ |
| Top 30% of distribution | Remaining 70% | 1.60 (0.77–3.42) | $2.11x10^{-01}$ |
| Top 20% of distribution | Remaining 80% | 2.20 (0.94–5.56) | $8.06x10^{-02}$ |
| Top 10% of distribution | Remaining 90% | 6.39 (1.65–42.63) | $1.89x10^{-02}$ |

**Supplementary Table 6. Comparison of Base and Full Cox Proportional Hazards Models for Predicting Time to Keratoplasty in the Validation Cohort**

| Model / Subgroup | Predictor | Hazard Ratio (95% CI) | P Value | Concordance (SE) |
|---|---|---|---|---|
| **Base model* (Overall)** | CTG18.1 expansion (Exp+) | 2.04 (1.22–3.41) | .006 | 0.679 (0.026) |
| | Sex (male) | 0.93 (0.61–1.40) | .71 | |
| **Full model (Overall)** | CTG18.1 expansion (Exp+) | 1.70 (1.00–2.88) | .049 | 0.689 (0.026) |
| | PRS (per 1-SD increase) | 1.42 (1.15–1.75) | .001 | |
| | Sex (male) | 0.90 (0.59–1.35) | 0.6 | |
| Sensitivity Analysis | | | | |
| **Full model (CTG18.1 Exp+ only)** | PRS (per 1-SD increase) | 1.40 (1.11–1.75) | .005 | 0.680 (0.028) |
| | Sex (male) | 0.88 (0.56–1.39) | 0.57 | |
| **Full model (CTG18.1 Exp- only)** | PRS (per 1-SD increase) | 2.94 (1.13–7.68) | .0276 | 0.766 (0.069) |
| | Sex (male) | 0.32 (0.07–1.50) | .1497 | |
| *Base model includes CTG18.1 expansion, sex, and 10 genetic principal components (PCs). Full model additionally includes standardized polygenic risk score (PRS). | | | | |

#### Supplementary Figures

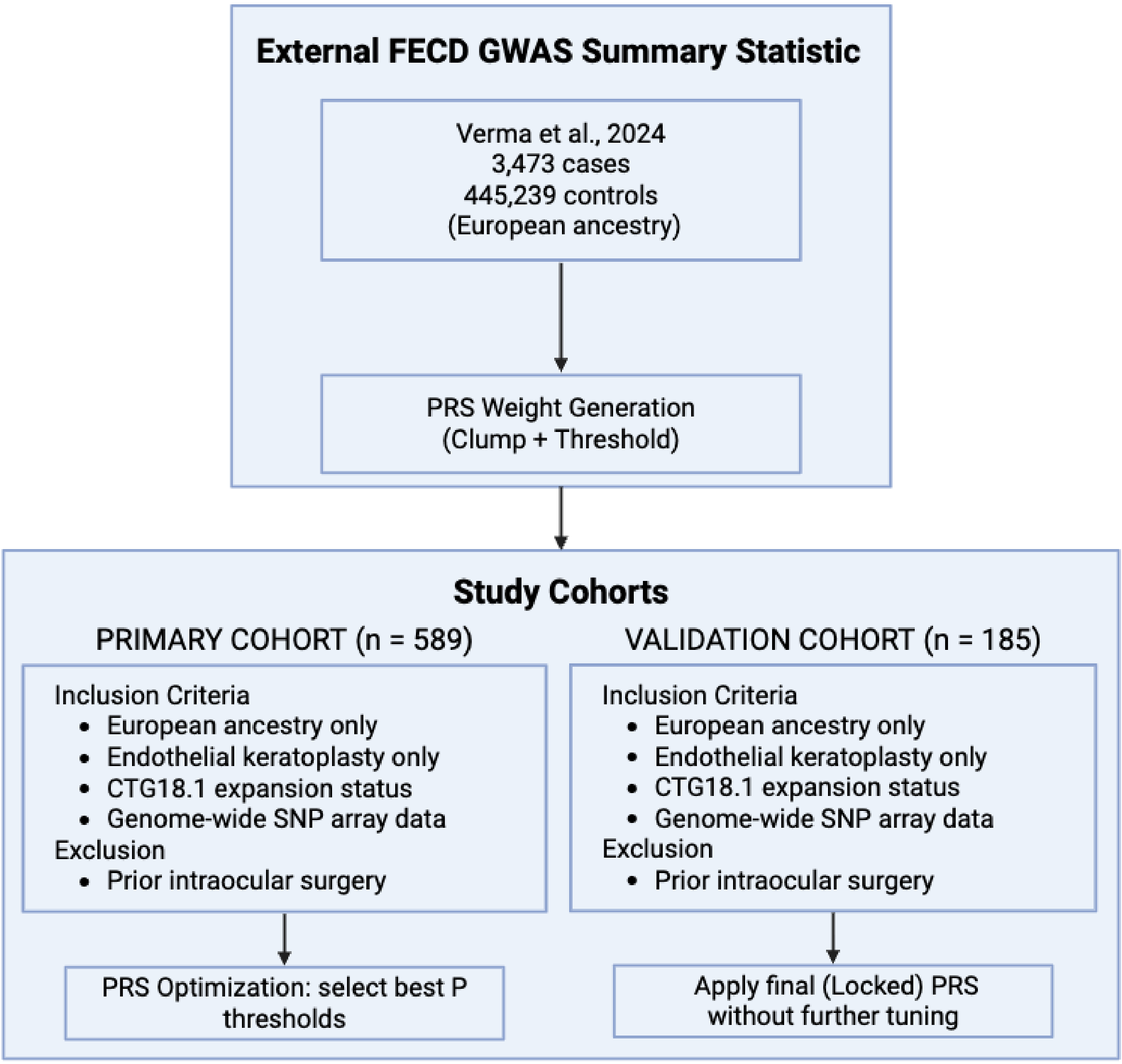


**Supplementary Figure 1. Overview of the Study Workflow**

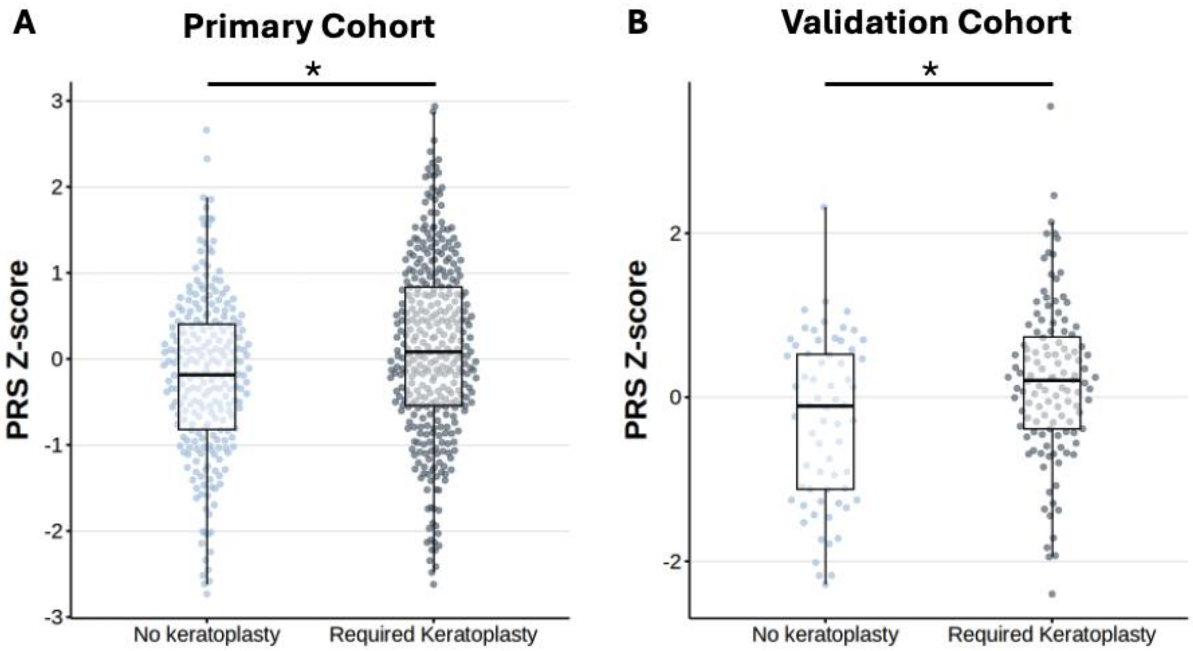


**Supplementary Figure 2. Polygenic Risk Score Distribution and Association with Keratoplasty Status.** Distribution of standardised polygenic risk score (PRS) among participants stratified by keratoplasty history in the primary and validation cohort. Participants requiring keratoplasty had significantly higher mean PRS than those who remained transplant-free in both the primary cohort (**A;** 0.12 ± 1.05 vs −0.20 ± 0.94; P < .001) and the validation cohort (**B**; 0.20 ± 0.95 vs −0.29 ± 1.01; P = .001).

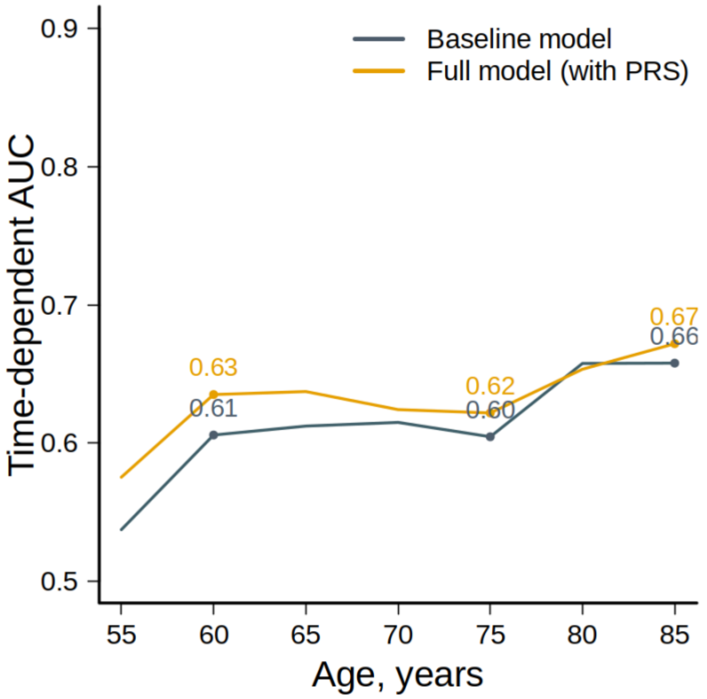


**Supplementary Figure 3. Inclusion of the Polygenic Risk Score Improves Prognostic Discrimination Over Time in the Primary Cohort.** Time-dependent area under the receiver operating characteristic curve (AUC[t]) for keratoplasty prediction models in the primary cohort. The full model outperformed the base model, demonstrating improved discrimination of individuals who would undergo keratoplasty (P = .03 by Wilcoxon signed-rank test).

*The base model included sex, the first 10 genetic principal components, and CTG18.1 expansion status. The full model additionally incorporated the standardized polygenic risk score (PRS).
